# ICU-level variation in arterial blood gas utilization and patient in-hospital mortality in critically ill patients: A retrospective cohort study using the Japanese Intensive care PAtient Database registry

**DOI:** 10.64898/2026.02.04.26345574

**Authors:** Shunsuke Yawata, Shigehiko Uchino, Seiichi Yamashima, Seiya Nishiyama, Shohei Ono, Yusuke Sasabuchi, Shinshu Katayama

## Abstract

**Background:** The role of arterial blood gas (ABG) testing in the intensive care unit (ICU) remains debated within the “less is more” paradigm. While unnecessary testing may pose risks without benefit, timely ABGs provide critical information in unstable patients. Institutional variation in early ABG utilization and its association with outcomes remains unclear.

**Methods:** We conducted a multicenter retrospective cohort study using the Japanese Intensive Care PAtient Database (JIPAD) between April 2015 and March 2023. Adult ICU patients with a stay ≥24 h and arterial line placement were included. The standardized number of ABGs (SNABGs) within the first 24 h was calculated as the ratio of observed to expected values, where expectations were derived from a multivariable model adjusting for patient covariates. ICUs were categorized into tertiles according to SNABG utilization. The primary outcome was in-hospital mortality, analyzed using multilevel logistic regression with ICU-level random intercepts. Restricted cubic splines were used to explore non-linear associations.

**Results:** Among 117,546 patients from 87 ICUs, the mean number of ABGs varied widely. After standardization, SNABGs ranged from 0.73–0.90 in the low tertile to 1.09–1.15 in the high tertile. In the multilevel model, SNABG was not significantly associated with in-hospital mortality (adjusted OR 0.942 [95% CI 0.807–1.100] for tertile 2; 0.874 [95% CI 0.751–1.017] for tertile 3). Flexible modeling suggested a non-linear trend toward better outcomes with higher utilization, but confidence intervals included unity.

**Conclusion:** Early ABG utilization varied across ICUs, yet was not significantly associated with mortality. Sensitivity analysis suggested a non-linear relationship, with a tendency toward better outcomes at higher utilization. These findings warrant further investigation to clarify the role of early ABG utilization in critical care.

## Introduction

”Less is more” is the concept that overuse of medical care may result in harm, whereas less care can sometimes lead to better health outcomes. In critical care, this principle has been demonstrated not only for therapeutic interventions[1–3], but also for diagnostic procedures[4–6]. On the other hand, in the acute phase, such interventions and procedures may not necessarily conform to the “less is more” paradigm[7].

Arterial blood gas measurements (ABGs) provide comprehensive physiological information; however, complications associated with arterial line placement have been reported[8–10], and their routine use has often been questioned within the framework of the ”less is more” paradigm[11–15]. While unnecessary or frequent testing may expose patients to risks without clear benefits, timely ABGs in acute critical illness can play a crucial role in guiding therapeutic decisions[7]. This apparent tension suggests that “*less is not always more*” and highlights the need to clarify the appropriate role of ABGs in critical care. However, no studies have directly investigated the association between the frequency of ABGs and patient outcomes, particularly using a standardized measure of utilization.

In addition, clinical experience suggests that the frequency of ABG utilization varies substantially across ICUs. Although ICU-level variation in the total number of laboratory blood samples has been documented[16], whether such variation applies specifically to ABG testing and whether it affects patient outcomes remain uncertain. Therefore, we conducted a multicenter retrospective study using the Japanese Intensive Care PAtient Database (JIPAD)[17] to examine ICU-level variability in the early ABG utilization and its association with patient outcomes.

## Materials and Methods

### Study design and setting

This study was a retrospective analysis of the Japanese Intensive Care PAtient Database (JIPAD)[17]. JIPAD is a multicenter, prospective observational registry of critically ill patients, managed by the Japanese Society of Intensive Care Medicine. Established in 2014, JIPAD included 101 participating ICUs as of March 2023. In April 2018, with the launch of version 3.0, several data items (catecholamine use, high-flow nasal cannula (HFNC) use, platelet count, lactate, and sequential organ failure assessment (SOFA) score^10^) were added as data elements. Because of the anonymous nature of the data, the requirement for informed consent was waived. This study received approval from the Institutional Review Board at Jichi Medical University Saitama Medical Center (S24-809) on December 26, 2024.

### Participants

In the present study, the source population was JIPAD patients from April 2015 to March 2023. Inclusion criteria were: 1) age 18 years or older, 2) ICU length of stay of equal to or more than 24 hours, and 3) arterial line was placed. The exclusion criterion was the presence of missing data required for multivariable analysis.

### Outcome Measures and Data Collection

Variables collected from JIPAD included demographic data (age, sex, and body mass index [BMI]), admission-related factors (ICU admission source, emergency call activation, and hospital-to-ICU interval) and comorbidities (heart failure, respiratory failure, liver cirrhosis, metastatic cancer, immunosuppression, and maintenance dialysis), and primary diagnosis. Clinical data included the presence of a central venous catheter (CVC), mechanical ventilation (MV) within 24 hours of ICU admission, acute kidney injury (AKI) within 24 hours, infection, SOFA score, and APACHE III score[18]. Data on arterial blood gas analysis measurements (ABGs) and lactate levels within the first 24 hours were also collected. The JIPAD registry does not record the frequency of ABG measurements directly; instead, up to six sets of blood gas data (including FiO_2_, pH, PaO_2_, and PaCO_2_) are documented per patient. If more than six measurements are obtained, the six worst values are recorded according to predefined criteria. Lactate values were collected separately, with only the worst value recorded within 24 hours. Length of stay in the ICU and the hospital, and outcomes at ICU and hospital discharge were obtained from admission and discharge dates and survival status. ICU-level data were also obtained, including the number of hospital beds, ICU beds, intensivists, and ICU nurses, as well as hospital academic status. We additionally calculated the intensivist-to-ICU bed ratio and ICU nurse-to-ICU bed ratio as indicators of ICU staffing.

### Statistical Analysis

The primary exposure was the number of ABGs performed within the first 24 hours of ICU admission. Categorical variables were presented as counts and percentages, and continuous variables as medians with interquartile ranges (IQRs). Continuous variables including lactate, PaO_2_/FiO_2_ (P/F) ratio, PaCO_2_, and pH were categorized using clinically meaningful thresholds to enhance interpretability in regression models. The threshold for the P/F ratio was based on the SOFA score[19] while the cutoff for lactate was derived from literature related to SOFA and critical care outcomes[20]. Specifically, lactate was grouped into <2, 2–<4, 4–<6, 6–<10, and ≥10 mmol/L; Lowest P/F ratio into ≥400, 300–399, 200–299, 100–199, and <100 mmHg; Highest PaCO_2_ into <35, 35–45, >45–55, >55–60, and >60 mmHg; and Lowest pH into <7.15, 7.15–7.25, >7.25–7.35, >7.35–7.45, and >7.45. For blood gas data, an additional category labeled “Not measured” was created to account for patients without recorded values. For all other variables, missing data were handled using complete-case analysis; patients with missing values in any variable included in the multivariable model were excluded. The overall proportion of missing data was low, due to automated checks implemented in JIPAD at the time of data entry. Most of the missing values were for BMI. To account for differences in patient characteristics across ICUs, the expected number of ABGs was estimated using a multiple linear regression model based on patient-level covariates. We evaluated the calibration of the prediction model by dividing patients into deciles based on the predicted values, and plotting the mean predicted and observed numbers of ABGs within each decile. The standardized number of ABGs (SNABGs) was then calculated as the ratio of the observed to expected values. We standardized ABG utilization to distinguish ICU-level practice patterns from differences driven by patient severity and case mix. ICUs were grouped into tertiles (low, medium, high utilization) based on their SNABGs for subsequent analyses. Differences across tertiles of SNABGs were assessed using the chi-square test for categorical variables and the Kruskal–Wallis test for continuous variables.

For the primary analysis, a multilevel logistic regression model with random intercepts for ICUs was used to examine the association between SNABGs and in-hospital mortality. This model accounted for clustering of patients within ICUs. To examine potential non-linear relationships, the association was modeled using restricted cubic splines with three degrees of freedom, allowing flexible yet interpretable estimation. Covariates were selected a priori based on clinical relevance and previous studies, and included demographic characteristics, comorbidities, severity of illness, ICU admission source, and ICU-level characteristics such as staffing. For interpretability, odds ratios were centered at SNABG = 1.0, representing the expected number of ABG measurements per patient. While ICU-level random intercepts were included to account for unmeasured baseline differences in mortality across ICUs, SNABG was treated as an ICU-level exposure of interest, reflecting differences in utilization practices rather than residual confounding.

All analyses were performed using R software (version 4.4.2, R Foundation for Statistical Computing, Vienna, Austria). A two-sided p-value of <0.05 was considered statistically significant. The analysis code is provided as Supporting Information (S1 File).

## Results

### Study flow

A total of 324,396 patients were registered in JIPAD from April 2015 to March 2023 across 101 participating ICUs. Of these, 21,303 patients were excluded due to missing ICU-level data from 8 ICUs (n = 13,972) or registration at pediatric ICUs (5 ICUs, n = 7,331), leaving 303,093 patients from 88 ICUs. An additional 54,553 patients were excluded because their data were collected before the implementation of JIPAD version 3.0, resulting in 248,540 patients eligible for further screening. Among these, 130,994 patients were excluded for the following reasons: age under 16 years (n = 8,462), ICU stay of less than 24 hours (n = 106,031), absence of an arterial line (n = 7,928), ICU readmission (n = 7,700), and missing data on outcomes (n = 1) or body mass index (n = 872). Finally, 117,546 patients from 87 ICUs were included in the analysis (Fig 1).

**Fig 1.**
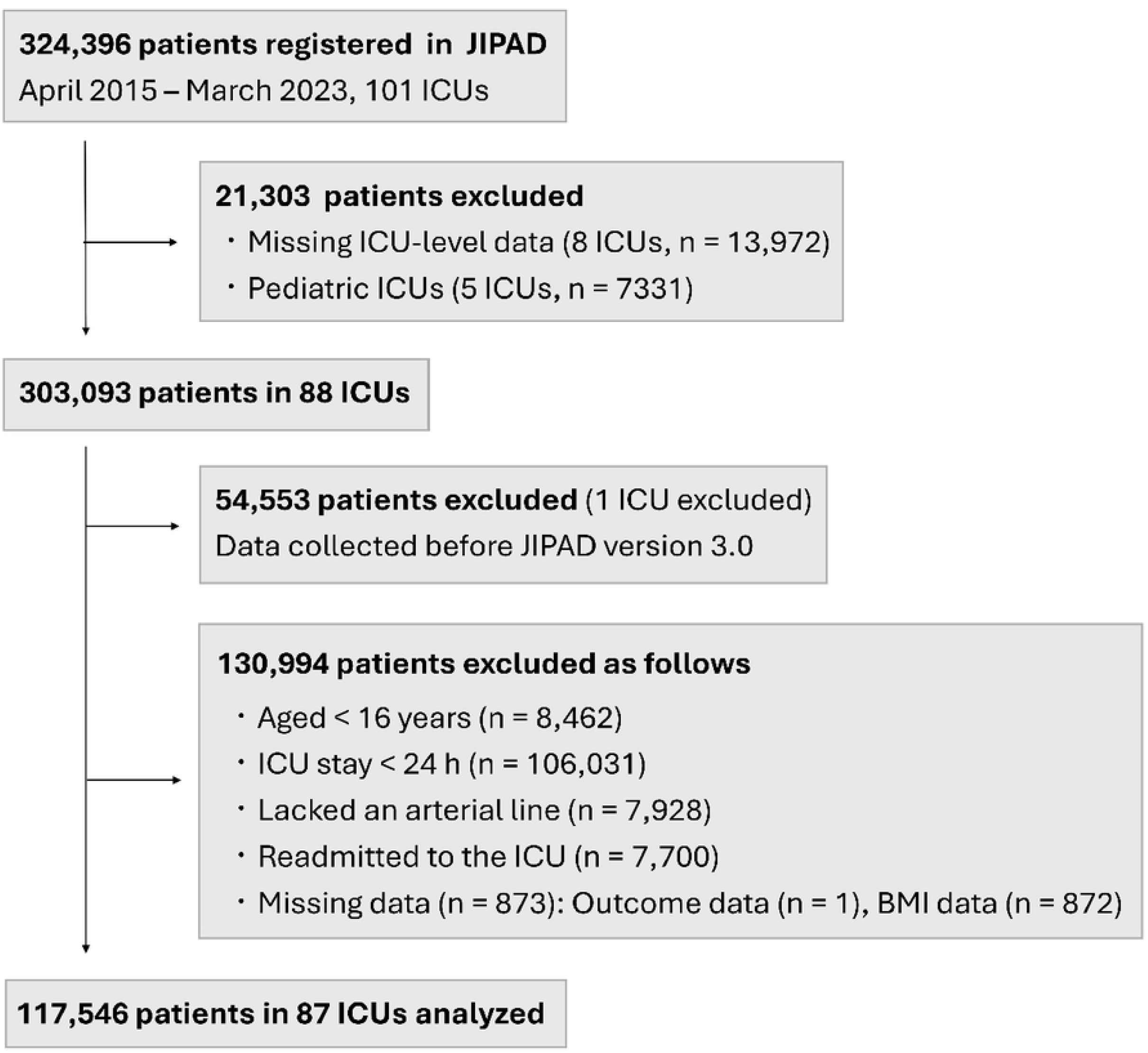
Patient flow. Selection of adult ICU admissions from the JIPAD database between April 2015 and March 2023. Abbreviations: ICU, intensive care unit; BMI, body mass index; JIPAD, Japanese Intensive Care Patient Database.

### ICU-Level Variation in Arterial Blood Gas Utilization

The frequency of ABGs varied substantially across ICUs. Fig 2 shows the average number of ABGs per ICU, with ICUs sorted in ascending order of average ABGs utilization. As shown in the Figure, the number of ABGs—recorded up to a maximum of six within the first 24 hours in JIPAD— averaged approximately 2 times in low-utilization ICUs, and nearly 6 times in high-utilization ICUs.

**Fig 2.**
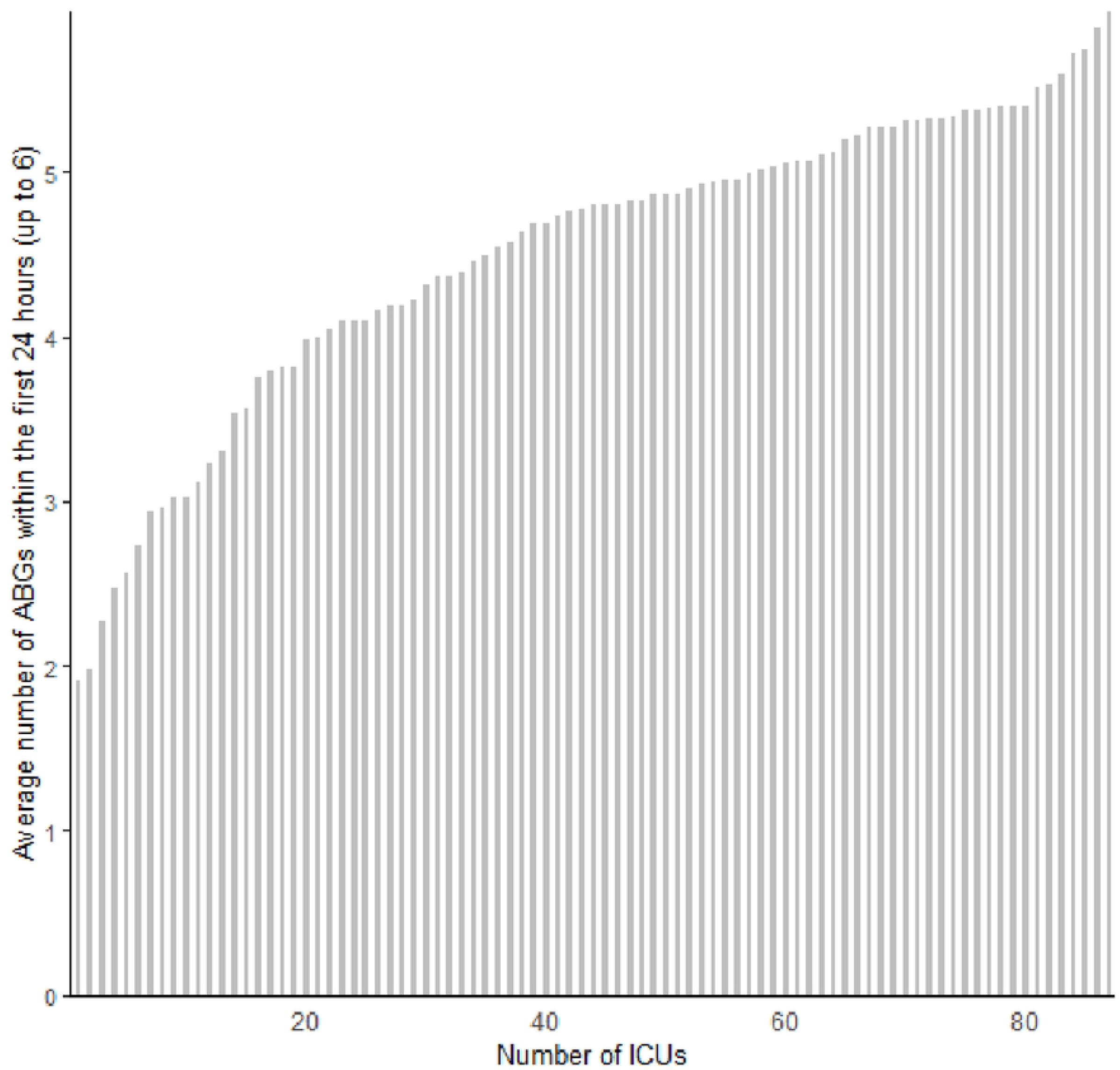
Variation in Arterial Blood Gas Analysis Measurement Frequency Across ICUs (Sorted by Lowest to Highest). Mean number of ABGs per patient within the first 24 hours after ICU admission for each of the 87 ICUs, ordered from lowest to highest. ABGs were recorded up to a maximum of six within the first 24 hours in JIPAD. Abbreviations: ABGs, Arterial Blood Gas Analysis Measurement; JIPAD, Japanese Intensive Care Patient Database.

### Standardized ABGs Utilization and Classification

We calculated the standardized number of ABGs (SNABGs) using the observed and expected numbers of ABGs, where the expected number was estimated by a multiple linear regression model based on patient-level covariates (S1 Table). The calibration plot demonstrated that the predicted and observed numbers of ABGs were in good agreement across deciles (Fig 3).

**Fig 3.**
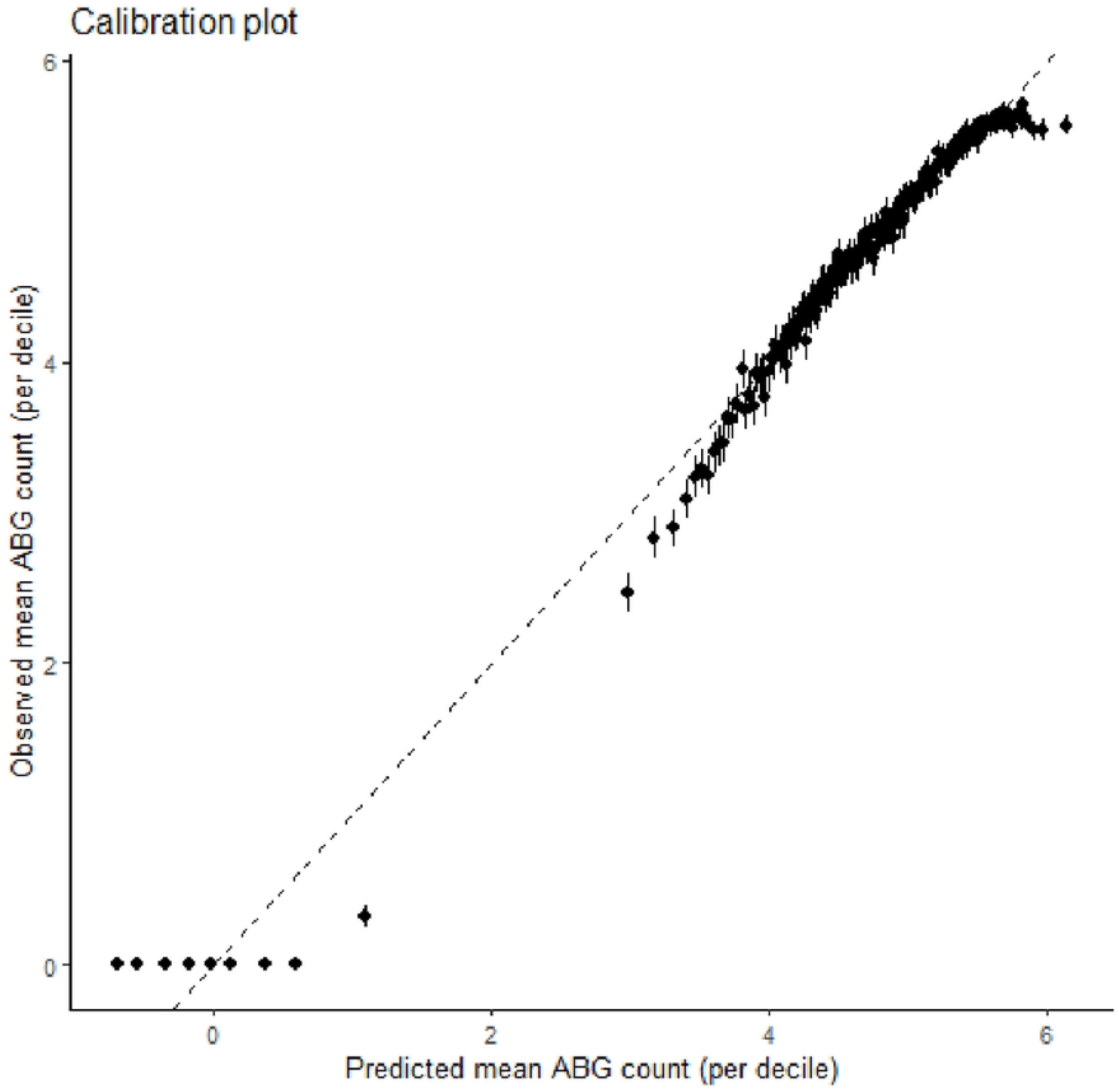
Calibration of the prediction model for expected number of ABGs. Calibration plot showing the relationship between predicted and observed numbers of ABGs within the first 24 hours after ICU admission. Patients were grouped into deciles based on predicted values from the multiple linear regression model. The x-axis represents the mean predicted number of ABGs within each decile, and the y-axis represents the mean observed number of ABGs. The diagonal line indicates perfect calibration. Abbreviations: ABGs, Arterial Blood Gas Analysis Measurement

Fig 4 shows the relationship between predicted and actual numbers of ABGs per ICU. ICUs in the high tertile generally had observed values exceeding predictions, whereas those in the low tertile had fewer ABGs than expected given patient characteristics. Tertile 1 had a mean SNABGs of 0.83 [Interquartile Range (IQR): 0.73–0.90], tertile 2 had 1.01 [IQR: 0.98–1.04], and tertile 3 had 1.11 [IQR: 1.09–1.15].

**Fig 4.**
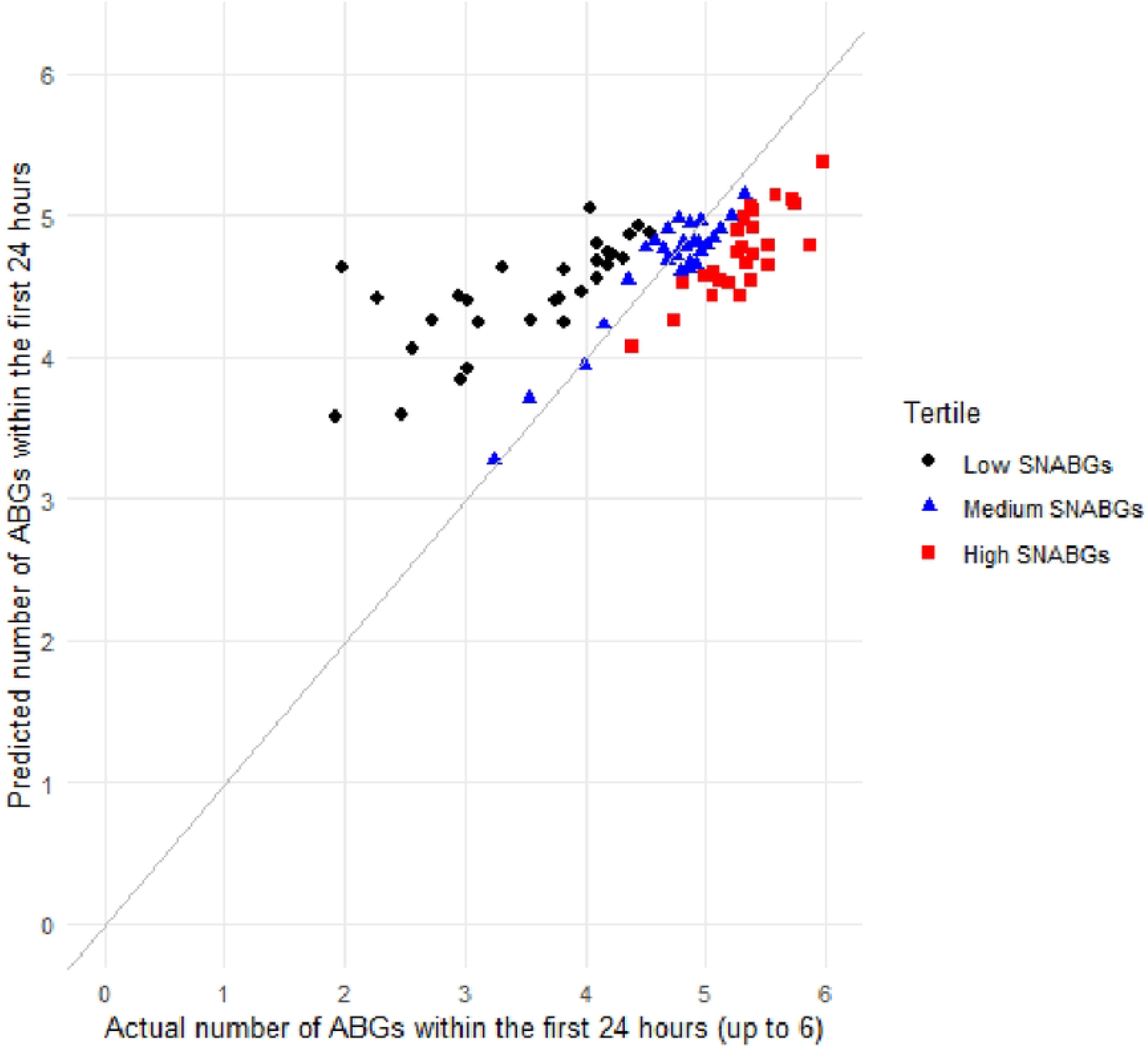
Predicted vs. Actual Number of Arterial Blood Gas Analysis per ICU, Colored by Standardized Utilization Tertile. Each point represents one ICU; the diagonal line denotes equality (predicted = observed). Predicted values are from a multivariable model adjusting for patient characteristics; ICUs are colored by tertiles of SNABGs. ICUs are shown as black circles (low SNABGs tertile), blue triangles (medium tertile), and red squares (high tertile). Abbreviations: ABGs, Arterial Blood Gas Analysis Measurement; SNABGs, Standardized number of ABGs

The characteristics of patients are summarized in Table 1. Although several variables differed significantly across tertiles, the absolute differences were modest in size. ICU-level characteristics across the tertiles of standardized ABG utilization are shown in Table 2. While the number of patients per ICU tended to increase with higher ABG utilization, other ICU characteristics such as the number of hospital beds, ICU beds, and staffing levels did not show statistically significant differences across tertiles. ICU mortality was significantly lower in the high-utilization tertile compared to the lower tertiles, whereas in-hospital mortality and lengths of stay were comparable across groups.

**Table 1.**
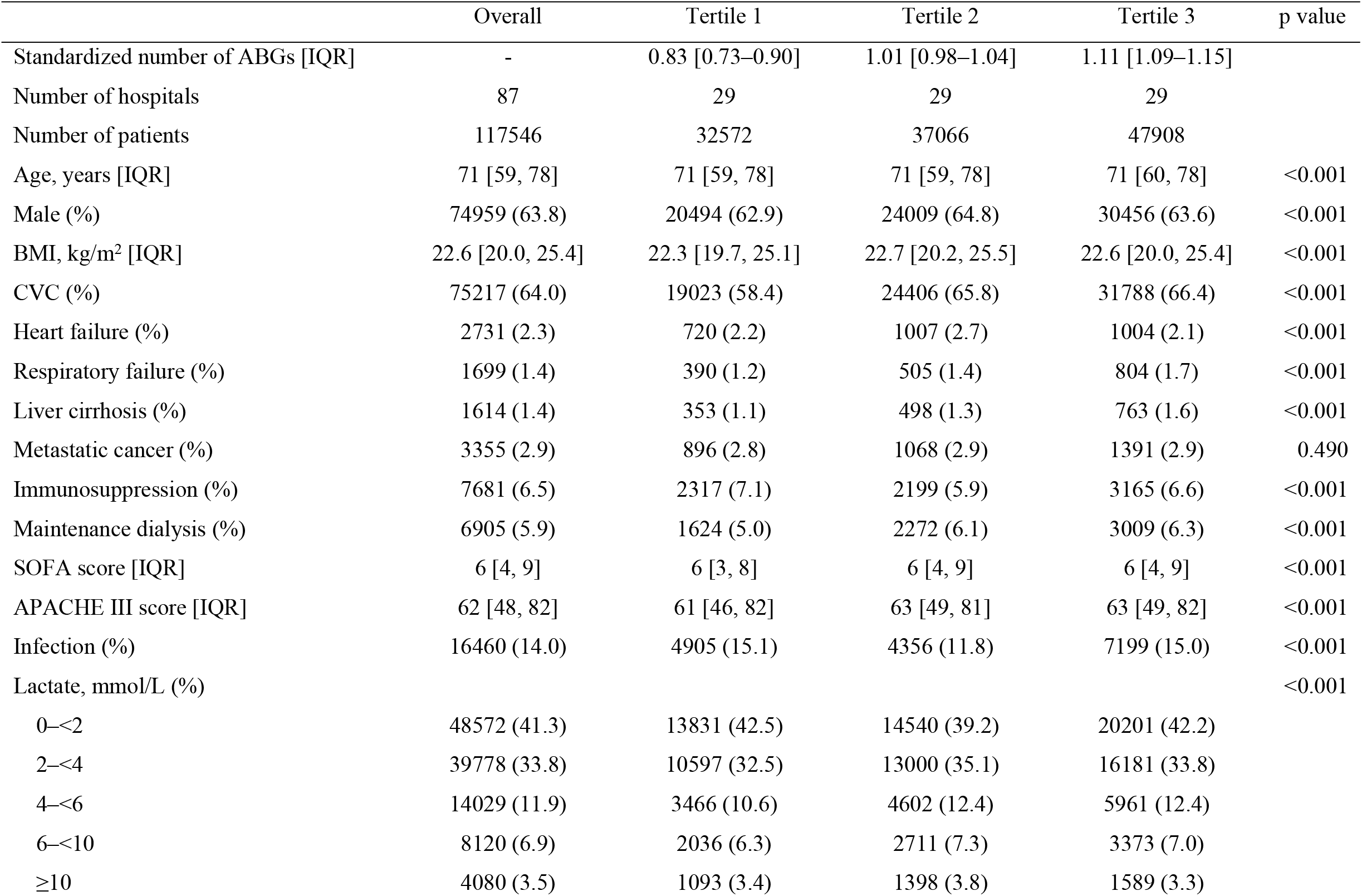
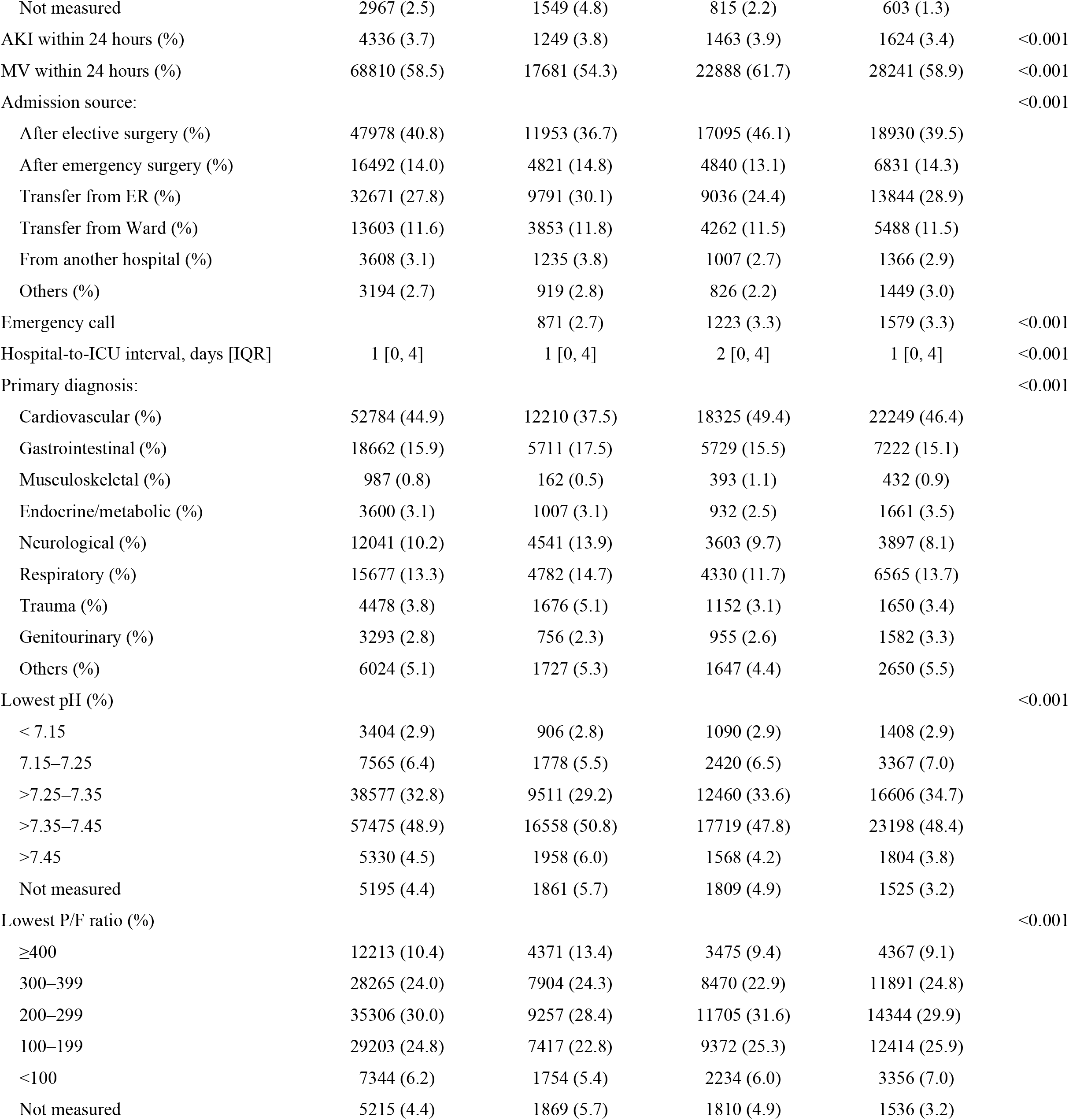
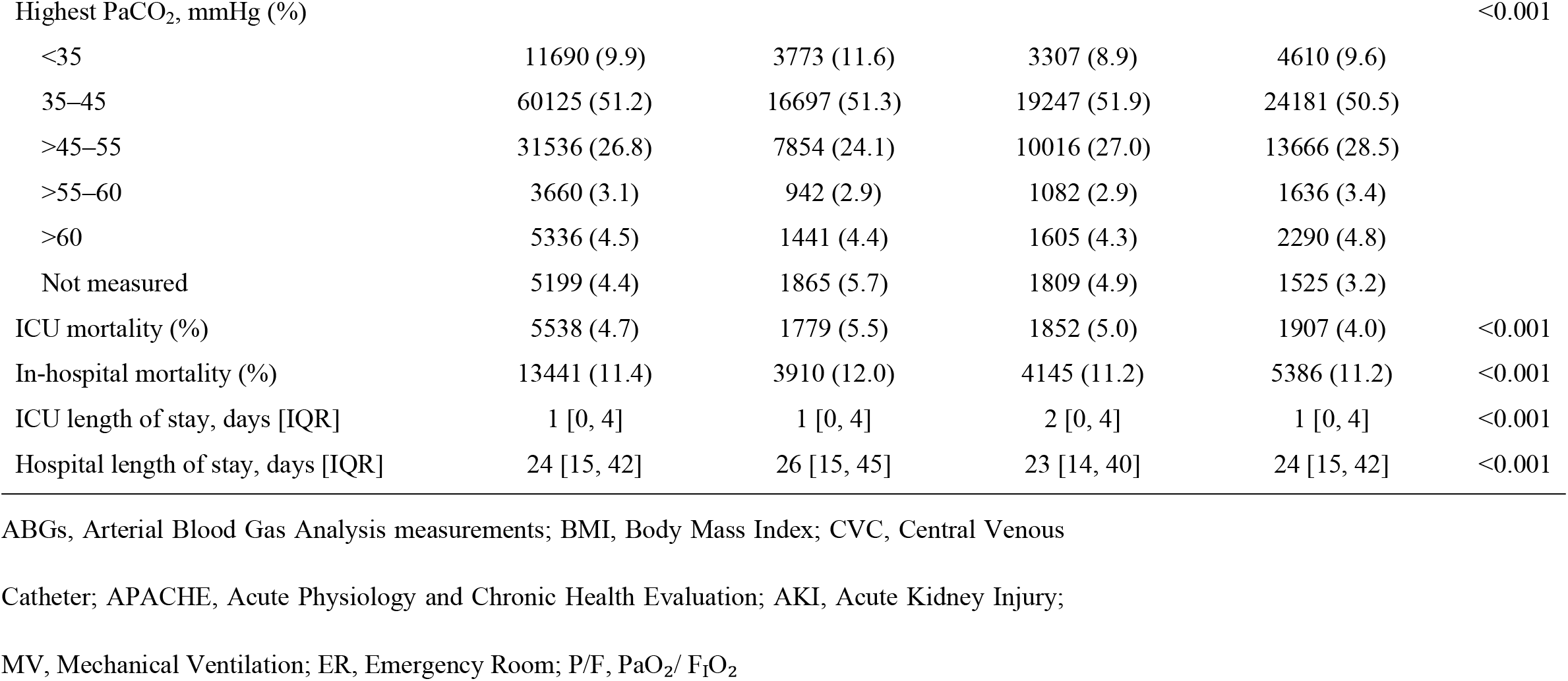
Patient Characteristics.

**Table 2.**
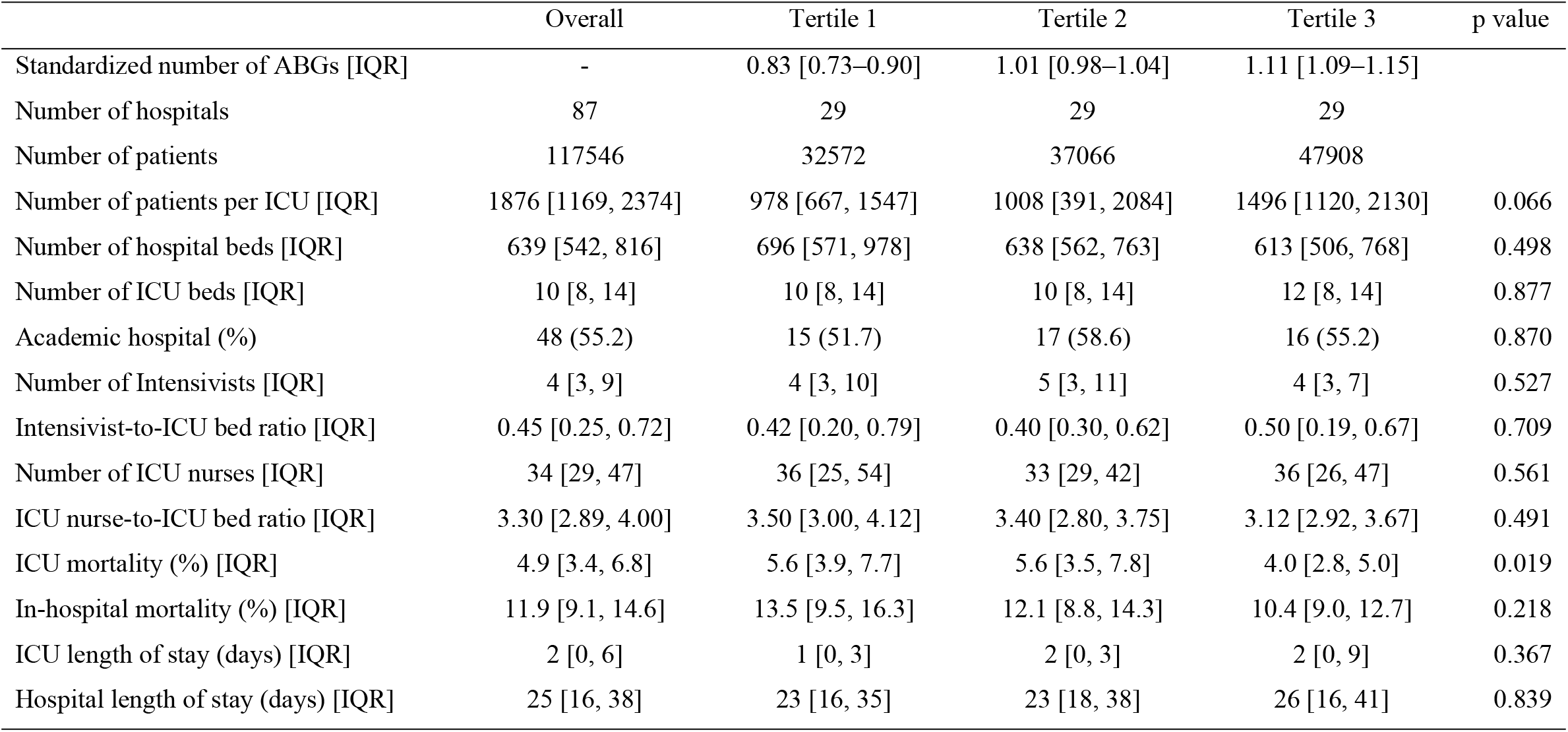
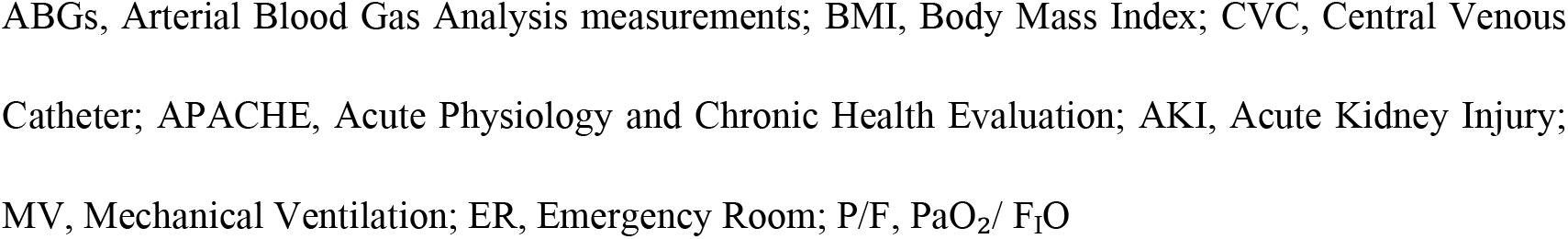
ICU-Level Characteristics.

### Association Between SNABG Utilization and In-Hospital Mortality

In the multilevel logistic regression model with random intercepts for ICUs, SNABG utilization was not significantly associated with in-hospital mortality (Table 3). The adjusted odds ratio was 0.942 (95% CI: 0.807–1.100) for tertile 2 and 0.874 (95% CI: 0.751–1.017) for tertile 3. To further examine the association, SNABG was modeled as a continuous variable using restricted cubic splines. This analysis suggested a non-linear association with in-hospital mortality (Fig 5): the adjusted odds ratio increased slightly below the median SNABG level and decreased at higher levels, suggesting a downward-sloping curve beyond the average value. However, the 95% confidence interval included 1.0 across much of the range.

**Table 3.**
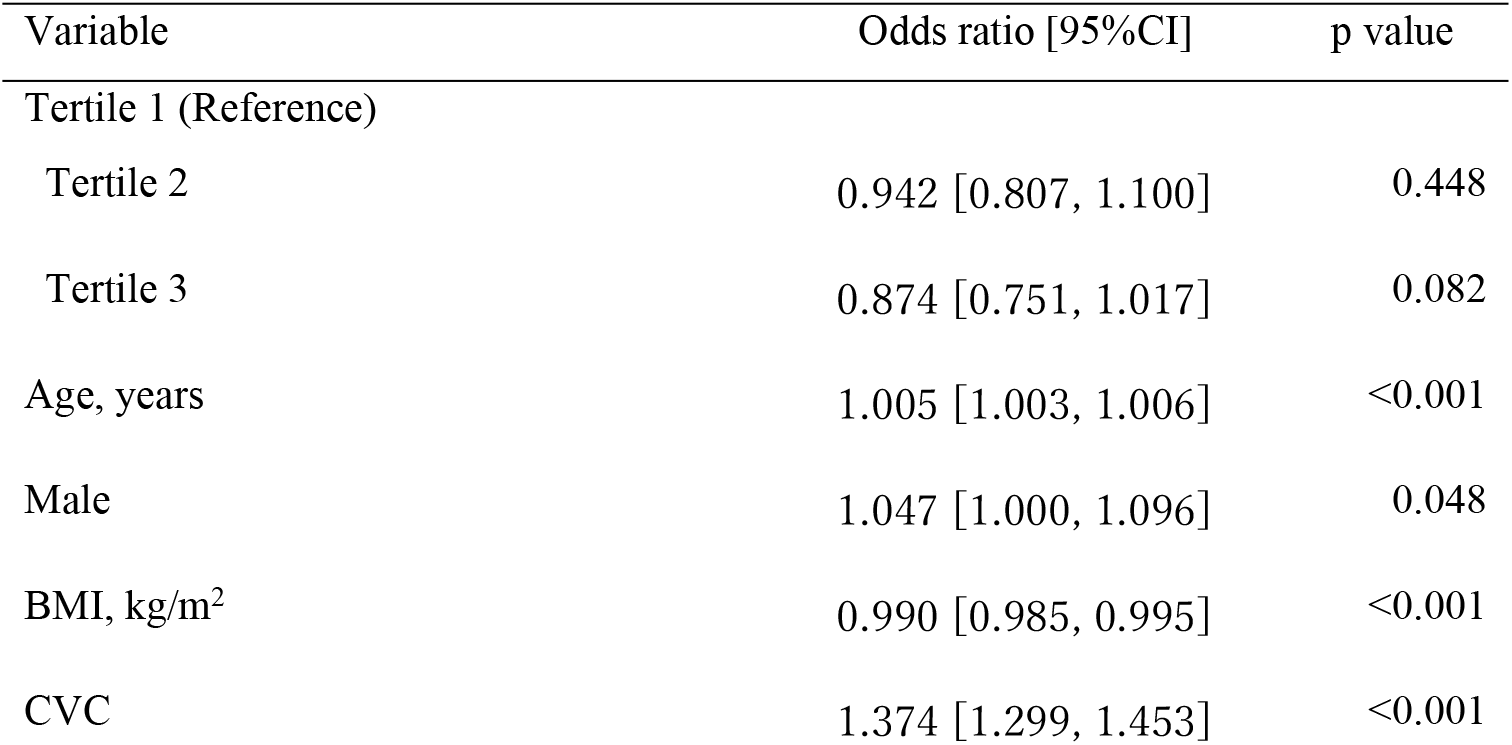
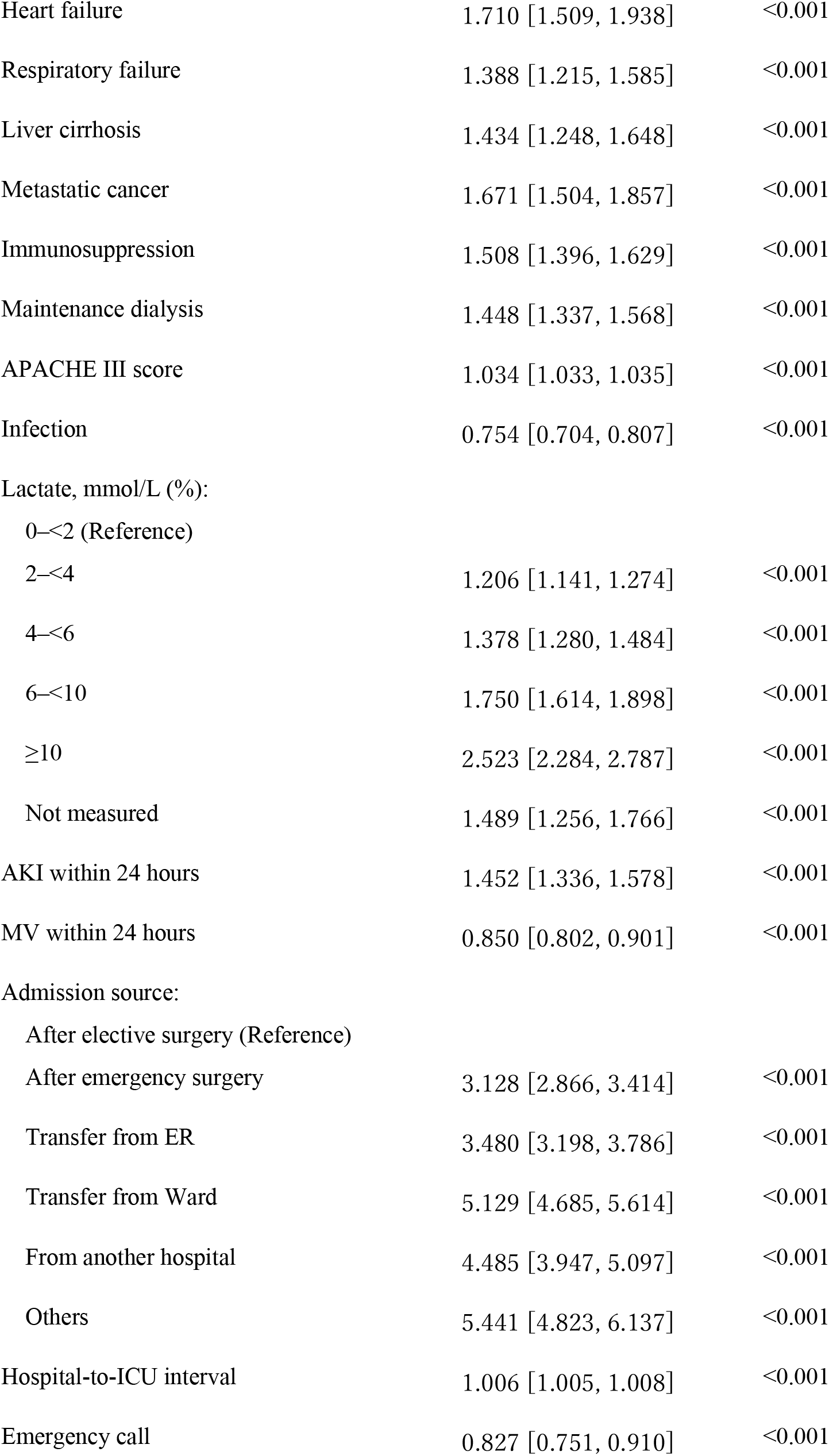
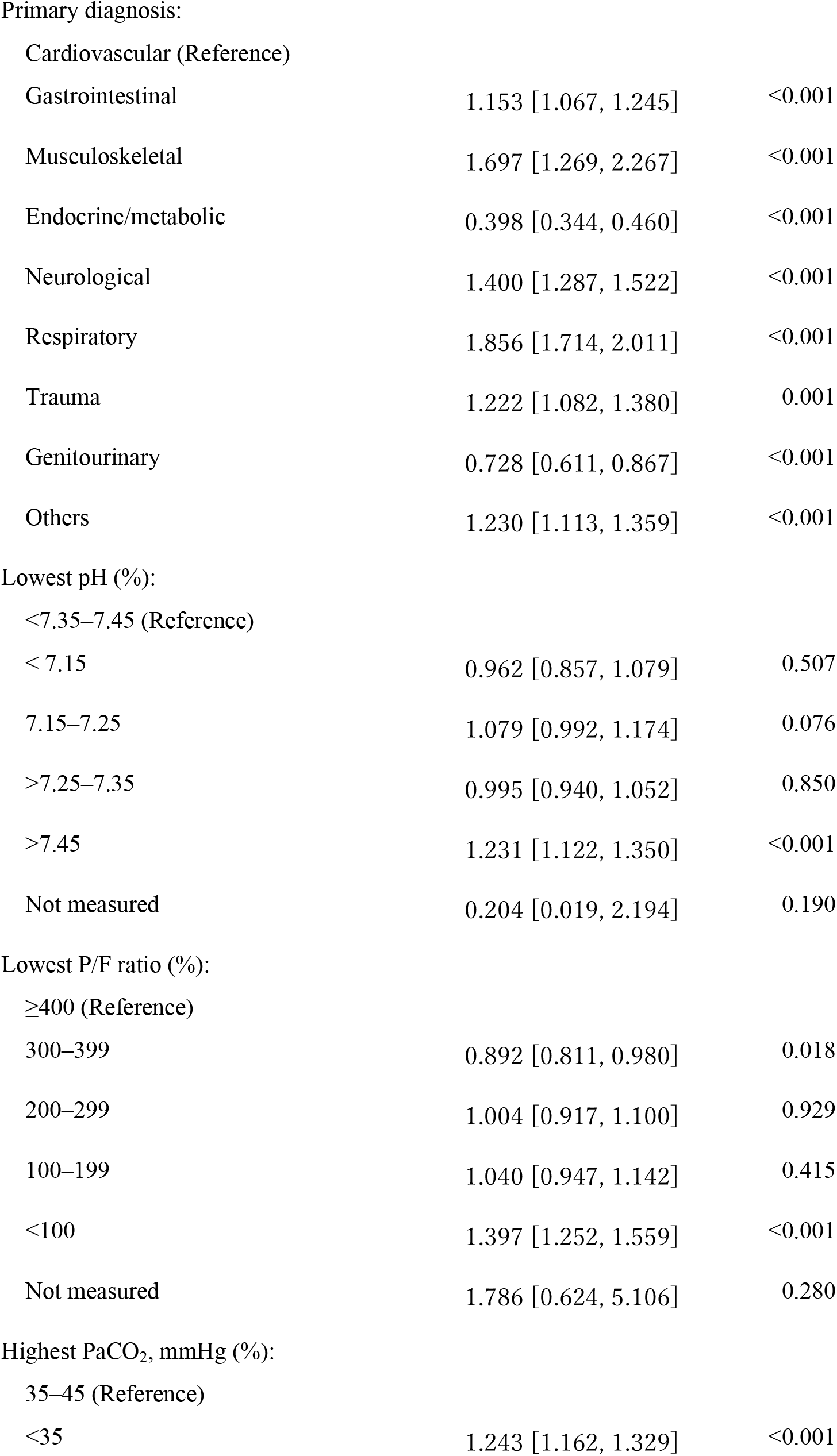
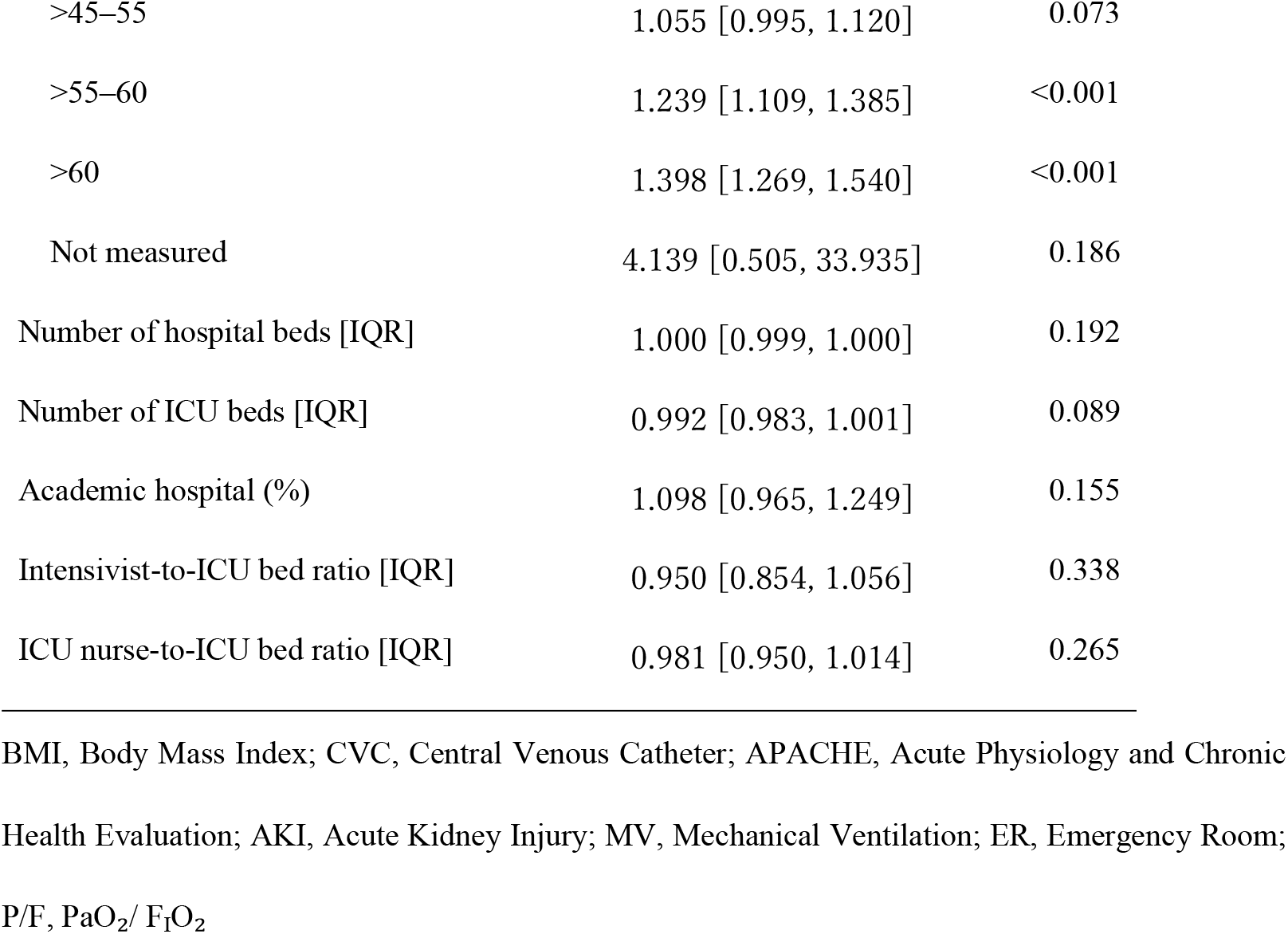
Multilevel Logistic Regression Model for Arterial Blood Gas Analysis Measurement.

| Variable | Odds ratio [95%CI] | p value |
| --- | --- | --- |
| Tertile 1 (Reference) |  |  |
| Tertile 2 | 0.942 [0.807, 1.100] | 0.448 |
| Tertile 3 | 0.874 [0.751, 1.017] | 0.082 |
| Age, years | 1.005 [1.003, 1.006] | <0.001 |
| Male | 1.047 [1.000, 1.096] | 0.048 |
| BMI, kg/m<sup>2</sup> | 0.990 [0.985, 0.995] | <0.001 |
| CVC | 1.374 [1.299, 1.453] | <0.001 |
| Heart failure | 1.710 [1.509, 1.938] | <0.001 |
| Respiratory failure | 1.388 [1.215, 1.585] | <0.001 |
| Liver cirrhosis | 1.434 [1.248, 1.648] | <0.001 |
| Metastatic cancer | 1.671 [1.504, 1.857] | <0.001 |
| Immunosuppression | 1.508 [1.396, 1.629] | <0.001 |
| Maintenance dialysis | 1.448 [1.337, 1.568] | <0.001 |
| APACHE III score | 1.034 [1.033, 1.035] | <0.001 |
| Infection | 0.754 [0.704, 0.807] | <0.001 |
| Lactate, mmol/L (%): |  |  |
| 0-<2 (Reference) |  |  |
| 2-<4 | 1.206 [1.141, 1.274] | <0.001 |
| 4-<6 | 1.378 [1.280, 1.484] | <0.001 |
| 6-<10 | 1.750 [1.614, 1.898] | <0.001 |
| ≥10 | 2.523 [2.284, 2.787] | <0.001 |
| Not measured | 1.489 [1.256, 1.766] | <0.001 |
| AKI within 24 hours | 1.452 [1.336, 1.578] | <0.001 |
| MV within 24 hours | 0.850 [0.802, 0.901] | <0.001 |
| Admission source: |  |  |
| After elective surgery (Reference) |  |  |
| After emergency surgery | 3.128 [2.866, 3.414] | <0.001 |
| Transfer from ER | 3.480 [3.198, 3.786] | <0.001 |
| Transfer from Ward | 5.129 [4.685, 5.614] | <0.001 |
| From another hospital | 4.485 [3.947, 5.097] | <0.001 |
| Others | 5.441 [4.823, 6.137] | <0.001 |
| Hospital-to-ICU interval | 1.006 [1.005, 1.008] | <0.001 |
| Emergency call | 0.827 [0.751, 0.910] | <0.001 |
| Gastrointestinal | 1.153 [1.067, 1.245] | <0.001 |
| Musculoskeletal | 1.697 [1.269, 2.267] | <0.001 |
| Endocrine/metabolic | 0.398 [0.344, 0.460] | <0.001 |
| Neurological | 1.400 [1.287, 1.522] | <0.001 |
| Respiratory | 1.856 [1.714, 2.011] | <0.001 |
| Trauma | 1.222 [1.082, 1.380] | 0.001 |
| Genitourinary | 0.728 [0.611, 0.867] | <0.001 |
| Others | 1.230 [1.113, 1.359] | <0.001 |
| < 7.15 | 0.962 [0.857, 1.079] | 0.507 |
| 7.15–7.25 | 1.079 [0.992, 1.174] | 0.076 |
| >7.25–7.35 | 0.995 [0.940, 1.052] | 0.850 |
| >7.45 | 1.231 [1.122, 1.350] | <0.001 |
| Not measured | 0.204 [0.019, 2.194] | 0.190 |
| 300–399 | 0.892 [0.811, 0.980] | 0.018 |
| 200–299 | 1.004 [0.917, 1.100] | 0.929 |
| 100–199 | 1.040 [0.947, 1.142] | 0.415 |
| <100 | 1.397 [1.252, 1.559] | <0.001 |
| Not measured | 1.786 [0.624, 5.106] | 0.280 |
| <35 | 1.243 [1.162, 1.329] | <0.001 |
| >45–55 | 1.055 [0.995, 1.120] | 0.073 |
| >55–60 | 1.239 [1.109, 1.385] | <0.001 |
| >60 | 1.398 [1.269, 1.540] | <0.001 |
| Not measured | 4.139 [0.505, 33.935] | 0.186 |
| Number of hospital beds [IQR] | 1.000 [0.999, 1.000] | 0.192 |
| Number of ICU beds [IQR] | 0.992 [0.983, 1.001] | 0.089 |
| Academic hospital (%) | 1.098 [0.965, 1.249] | 0.155 |
| Intensivist-to-ICU bed ratio [IQR] | 0.950 [0.854, 1.056] | 0.338 |
| ICU nurse-to-ICU bed ratio [IQR] | 0.981 [0.950, 1.014] | 0.265 |

BMI, Body Mass Index; CVC, Central Venous Catheter; APACHE, Acute Physiology and Chronic Health Evaluation; AKI, Acute Kidney Injury; MV, Mechanical Ventilation; ER, Emergency Room; P/F, PaO<sub>2</sub>/F<sub>I</sub>O<sub>2</sub>

**Fig 5.**
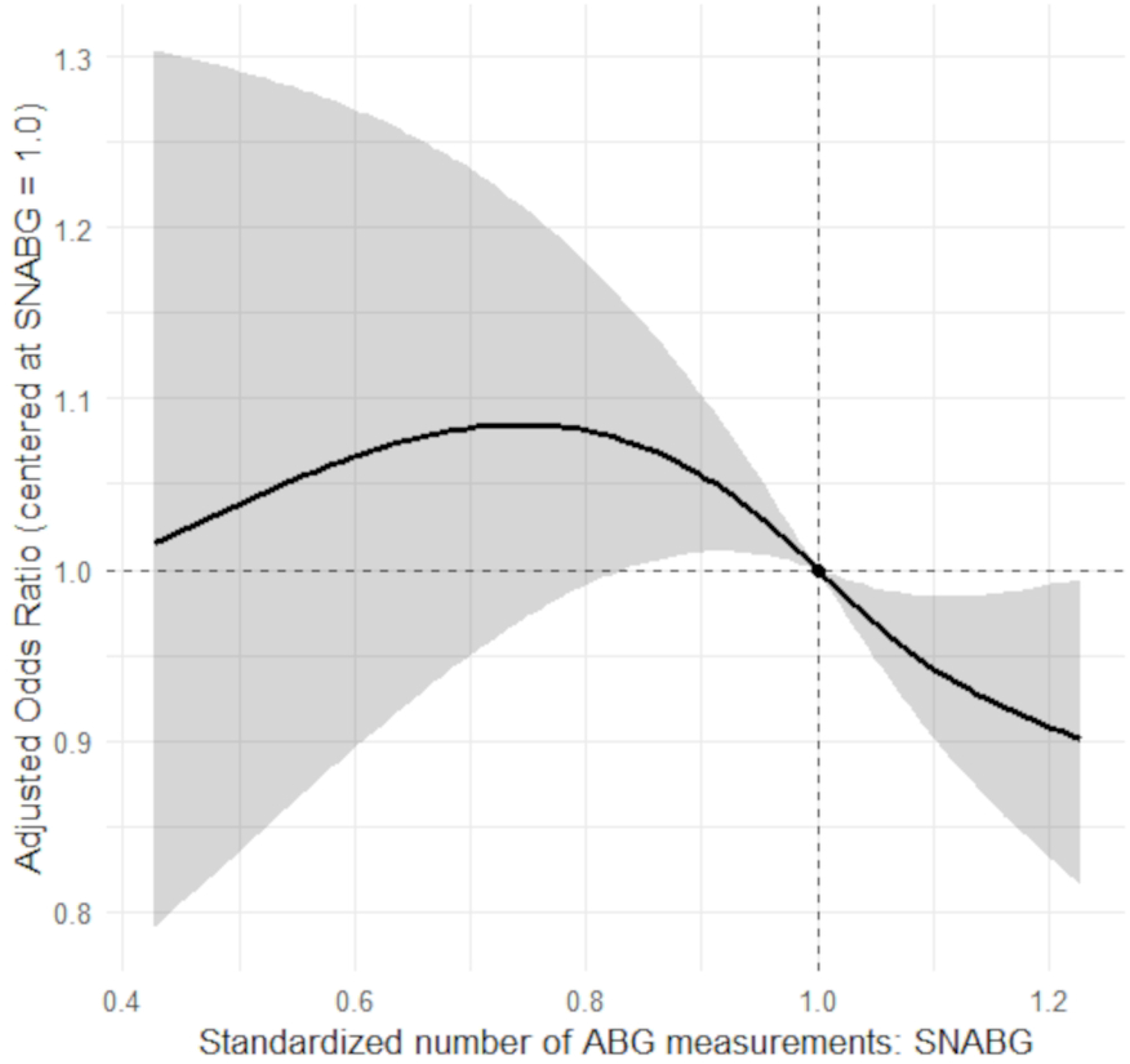
Association Between Standardized Number of Arterial Blood Gas Analysis Measurements and In-Hospital Mortality: Adjusted Odds Ratio Modeled Using Restricted Cubic Splines in a Multilevel Logistic Regression Model. Multilevel logistic regression with restricted cubic splines (df = 3), centered at SNABG = 1.0. The solid line shows the adjusted odds ratio; the shaded band shows the 95% confidence interval. Random intercepts account for clustering by ICU. Abbreviations: ABGs, Arterial Blood Gas Analysis Measurement; SNABGs, Standardized number of ABGs; OR, Odds Ratio

## Discussion

In this large multicenter study of more than 117,000 ICU patients, we found substantial ICU-level variation in early ABG utilization. However, standardized utilization was not significantly associated with in-hospital mortality. When examined using flexible modeling, the association suggested a potential non-linear trend toward better outcomes with higher utilization.

The concept of “less is more” has been increasingly interpreted not only in the narrow sense of “more is harmful and less is beneficial,” but also in the broader sense of “less is non-inferior and more is not necessarily beneficial.” In critical care, routine procedures have been questioned under this paradigm. For example, in a large multicenter RCT involving 1,041 patients considered for pulmonary artery catheterization, insertion was not associated with improved survival compared with no insertion (adjusted hazard ratio 1.09, 95% CI 0.94 – 1.27)[4]. Recent findings have similarly questioned whether routine arterial catheterization improves outcomes in shock. In the EVERDAC trial, early arterial catheterization did not reduce 28-day mortality (34.3% in the noninvasive-strategy group vs. 36.9% in the invasive-strategy group). The adjusted absolute risk difference was -3.2 percentage points (95% CI -8.9 – 2.5), confirming noninferiority. These findings reinforce the notion that monitoring tools do not inherently confer benefit unless the information they provide is effectively translated into clinical action[21]. Similarly, in a descriptive study of routine chest radiographs in the ICU, new abnormalities were detected in only 5.8% and treatment changes were made in only 2.2% of cases[5]. A recent systematic review comparing routine and on-demand blood sampling reported no significant differences in mortality across 28 studies, while several studies demonstrated shorter ICU stays and reduced red blood cell transfusion rates with on-demand strategies[22]. ABG testing has likewise been examined through this lens. An educational intervention aimed at reducing unnecessary ABGs resulted in a 41.5% reduction in ABG use, saving 49 L of blood, 39,432 USD, and 1,643 staff hours annually, without worsening mortality or ICU length of stay[11].

However, all these studies evaluated routine testing during the entire ICU stay. In the very acute phase, when patients are unstable, the relationship between testing intensity and outcomes may differ. For instance, in an emergency department study, patients undergoing 1 to 3 procedures after intubation had significantly higher adjusted mortality than those receiving 5 to 6 procedures (OR 4.25, 95% CI 1.15 – 15.75)[23], suggesting that more active evaluation can be linked to better outcomes. Although ABG testing is associated with risks such as catheter-related complications and iatrogenic anemia, it provides essential information on oxygenation, ventilation, acid-base balance, electrolytes, glucose, and lactate, making ABGs particularly valuable during the acute phase of critical illness. Nevertheless, no previous study has directly examined the association between early ABG utilization and patient outcomes. In clinical practice, the frequency of ABG utilization varies substantially across ICUs, but the impact of this variation on patient outcomes remains unclear. A previous study evaluating blood sampling practices beyond the second ICU day reported that sampling was more frequent in teaching hospitals and in certain geographic regions[16]. Accordingly, in this study, we used JIPAD to examine the association between ICU-level ABG utilization within the first 24 hours of ICU admission and in-hospital mortality.

Recent findings from the EVERDAC trial showed that early arterial catheterization in patients with shock did not improve clinical outcomes[21]. Although arterial catheters are often required to obtain ABGs, the lack of benefit from routine catheter placement does not directly inform the question of how frequently ABG data should be obtained or how this information is used. Importantly, previous studies have not evaluated whether variation in early ABG utilization itself is associated with patient outcomes, leaving uncertainty about the role of testing intensity during the initial phase of critical illness. Our study demonstrated wide ICU-level variation in ABG utilization within the first 24 hours of ICU admission, even after adjustment for patient-level characteristics. The multilevel analysis incorporating random intercepts for ICUs found no significant linear association between ABG frequency and in-hospital mortality. However, analyses allowing for non-linear associations suggested a potential trend toward better outcomes with higher utilization. This non-linear association is unlikely to be driven by ABG frequency alone and may instead reflect facility-level factors—such as the timeliness with which ABG information is translated into treatment decisions, the responsiveness of clinical teams, or the organizational culture surrounding physiological monitoring. Taken together, these findings suggest that reduced testing was not clearly harmful, yet more frequent testing was not uniformly beneficial. Rather than supporting a uniform “less is more” approach, they highlight the importance of context-dependent decision-making regarding ABG use in the acute phase of critical illness.

## Strengths and Limitations

To our knowledge, this is the first study to quantify ICU-level variation in early ABG utilization (within 24 hours of ICU admission) and to examine its association with patient outcomes. By introducing SNABGs and applying multilevel modeling with ICU-level random effects, we appropriately accounted for both patient- and ICU-level factors. Focusing on the first 24 hours of ICU admission—when patients are most unstable—adds clinical relevance. The inclusion of over 117,000 patients from 87 ICUs also enhances statistical precision and generalizability compared with single-center studies. In addition, the use of a national registry with standardized data collection strengthens the robustness of our findings.

However, this study also has several limitations. First, JIPAD records up to six ABG values per patient within 24 hours, prioritizing the most abnormal values. This approach likely underestimates true measurement frequency and may bias the dataset toward more abnormal values, particularly in high-utilization ICUs. As a result, the true impact of very frequent testing may not have been fully captured. Second, subsequent ABG measurement frequency could have been influenced by earlier results, raising the possibility of reverse causation. Nonetheless, institutional practices are often habitual, with some ICUs performing multiple ABGs routinely and others refraining even when abnormalities are present. The observed variation therefore likely reflects ICU-level practices rather than individual patient trajectories. Finally, JIPAD is a voluntary registry composed mainly of academic and tertiary centers, which may limit generalizability to community or non-tertiary ICUs, and may not fully reflect practice in other health care systems. Nevertheless, the large sample size and multicenter design provide valuable insights into ICU-level variation.

## Conclusions

This multicenter study of more than 117,000 patients from 87 ICUs examined the association between standardized ABG utilization within the first 24 hours of ICU admission and in-hospital mortality. ABG utilization varied substantially across ICUs, yet standardized use was not significantly associated with mortality. When modeled flexibly, the association did not follow a simple linear pattern and showed a tendency toward better outcomes at higher utilization levels. Lower utilization was not associated with worse outcomes, and higher utilization was not consistently associated with benefit. The observed non-linear trend warrants further investigation. Future research should address broader outcomes—including costs, resource utilization, and patient safety—through prospective high-quality designs to comprehensively evaluate the role of ABG utilization in critical care.

## Data Availability

The individual-level data from the Japanese Intensive Care Patient Database (JIPAD) are not publicly available due to data use agreements. The analysis code is provided as Supporting Information (S2 file.pdf).

## Supporting Information

**S1 Table. Multivariable Linear Regression Model for Predicting Arterial Blood Gas analysis Measurement**

BMI, Body Mass Index; CVC, Central Venous Catheter; APACHE, Acute Physiology and Chronic Health Evaluation; AKI, Acute Kidney Injury; MV, Mechanical Ventilation; ER, Emergency Room; P/F, PaO_2_/ F_I_O_2_

**S2 File. The analysis code**

